# Frequency of Aortic Miscannulation During Fluoroscopy-Free Endovascular Navigation

**DOI:** 10.64898/2025.11.28.25341214

**Authors:** Jason MacTaggart, Sruti Prathivadhi, Paul Aylward, Christian Sanderfer, Blake Marmie, Margarita Pipinos, Alicia Schiller, Alexey Kamenskiy

## Abstract

**Background:** Aortic wire cannulation is sometimes performed without the assistance of fluoroscopy for expediency, but the associated risk of miscannulation has not been quantified. We used human cadavers and an electronic endovascular simulator to characterize guidewire misplacement rates during attempted fluoroscopy-free cannulation of the descending thoracic aorta.

**Methods:** Aortic cannulation was done in perfused human cadavers (n=10, age 81±13 years) and in their vascular anatomies loaded into the Mentice VISTG5 simulator to validate the method. Trauma Computerized Tomography Angiography scans (n=89, age 47±24 years, range 5-93 years) were used to build 3D vascular models and assess morphometric variability in vessel tortuosity, length, and branch angulations. Aortic miscannulation frequency was measured using an electronic simulator with multiple passes using a wire-feeding mechanism, and the results were analyzed in the context of age.

**Results:** Aortic miscannulation was the same in cadavers as in the electronic simulator (12%) for all passes. Electronic simulation using a larger cohort demonstrated that miscannulations occurred in 36% of all subjects, and the frequency for all passes was 10±18%. Aortic miscannulation frequency was lower in <50-year-old subjects (5±11%) than in >50-year-old subjects (16±22%), and the most frequent misplacement locations were into the iliac (5%), celiac (2%), and the superior mesenteric arteries (1%).

**Conclusions:** Fluoroscopy-free aortic cannulation may be associated with significant miscannulation rates, particularly in subjects older than 50 years. Fluoroscopic confirmation of wire location or endovascular devices capable of detecting and avoiding aortic miscannulation may reduce aortic access complications.

## INTRODUCTION

Minimally invasive endovascular procedures have surpassed open surgery in treating a broad range of cardiovascular pathologies and trauma^1–6^. While vascular access through small incisions in peripheral arteries remote from the area of disease or injury often facilitates faster and safer treatment, it may be complicated by challenging vascular anatomies. In addition, life-saving procedures such as Resuscitative Endovascular Balloon Occlusion of the Aorta (REBOA) and Extracorporeal Membrane Oxygenation (ECMO) are increasingly performed out of the hospital and without fluoroscopy. In challenging tortuous anatomies, wire and device navigation can be difficult even with x-ray guidance, and fluoroscopy-free aortic miscannulation into smaller vessels and end organs can lead to complications even with ultrasound assistance^6^.

Assessment of aortic miscannulation in different vascular anatomies is not straightforward because blind insertion of wires and catheters in humans may be associated with clinical complications, and large animals typically have substantial anatomic variation from humans. Human cadaver models are a viable alternative, but they are usually limited to older subjects with tortuous anatomies^7,8^ and a higher prevalence of vascular disease. They are also expensive^9^, and the experiments are time-consuming, resulting in smaller sample sizes and potentially underpowered results. Electronic simulators allow the import of patient-specific anatomies and provide a relatively cost- and time-efficient way to evaluate miscannulation frequency in a large number of patients with a broad range of anatomies. However, these simulators were designed for medical education, and their fidelity in representing real navigation scenarios has not yet been comprehensively evaluated. In this study, we used human cadavers to validate one of the most popular electronic endovascular simulators, which was then used to assess aortic miscannulation frequency and final location during fluoroscopy-free navigation in a substantial number of human anatomies from a wide range of ages. We hypothesized that the electronic simulator would demonstrate similar results to physical human anatomies and that increasing age is associated with higher miscalculation rates.

## METHODS

### 2.1 Fluoroscopy-free aortic cannulation in human cadavers

A total of 10 cadavers (average age 81±13 years, range 54-98 years, 7 male, 3 female) lightly embalmed with glutaraldehyde-based solution^10^ and warm-perfused with radiopaque fluid containing calcium carbonate to avoid tissue swelling, were used to test the feasibility of fluoroscopy-free wire navigation to the middle of the descending thoracic aorta (aortic Zone I^11^ / aortic Zone 4^12^ by different classifications) using a regular stiffness regular angled guidewire (Terumo, Shibuya City, Tokyo, Japan). The wire was passed without the assistance of fluoroscopy through left and right percutaneous femoral access sites for a distance intended to reach the center of aortic Zone I^11^ based on torso surface measurements. A total of 3 wire passage attempts were made on each side, and the wire was removed entirely and inserted *de novo* for each attempt. A radiopaque ruler was affixed to each cadaver’s chest, and diluted contrast was injected into the aorta to verify the locations of its main branches. The end position of the wire was assessed with fluoroscopy and later verified by co-registering with 3D Computed Tomography Angiography (CTA) Imaging using the affixed ruler as a reference.

### 2.2 Three-dimensional reconstructions of cadaveric aortic anatomy

After performing wire navigation, each cadaver was taken to the clinical GE LightSpeed VCT XT 64-channel CTA scanner (GE Healthcare, Waukesha, USA), and the entire body was imaged with 0.625mm slice thickness and a resolution of 512 × 512 pixels. Three-dimensional reconstructions of the entire aorta and the common carotid, subclavian, axillary, celiac, superior mesenteric, renal, iliac, and common femoral arteries were performed using Mimics software (Materialize Co) and a combination of semi-automated thresholding, region growing, and volume separation techniques^7,8,13,14^. The resulting 3D vascular anatomies were exported as STL files and imported into the Mentice VIST G5 (Mentice AB, Gothenburg, Sweden) electronic endovascular simulator through the CASE-IT EVAR module.

### 2.3 Endovascular simulator calibration, validation of accuracy, and comparison of electronic measurements with human cadaver model results

The Mentice VIST G5 (Gothenburg, Sweden) electronic endovascular simulator was chosen because of its ability to import patient-specific anatomies and provide realistic wire advancement simulations. With the help of the manufacturer, simulator’s software was modified to produce 1-to-1 geometric scaling, and the navigation distances, accuracy, and repeatability were validated using phantom geometries (Figure A1 of the Appendix) constructed in Autodesk Inventor (San Rafael, California) and imported into VIST G5 in an STL format. These geometries allowed to test the simulator’s ability to register inserted wire lengths and evaluate how the wire traversed straight and curved anatomies at different advancement rates (Figure A2 of the Appendix).

Since VIST G5 uses optical tracking of the wire to measure insertion length, the speed of wire advancement can have a significant impact on the results. To account for this and ensure maximal accuracy and repeatability of measurements, a wire feeding system was developed (see additional details in the Supplement). Cadaver vascular anatomies reconstructed in Mimics were then loaded into the VIST G5 simulator, and each model was mapped to the template reference points to ensure the proper location of the vessels and other anatomic structures. The physical guidewire in VIST G5 was represented with a 0.035” 35° angle tip hydrophilic guidewire, and the wire insertion was done at 18.4 mm/s constant linear rate for a total of 3 trials for each side and each anatomy. Final locations of the guidewire tip were recorded, and the percentage of attempts that resulted in the wire tip ending in a small artery was calculated and compared with that observed in cadavers.

### 2.4 Three-dimensional reconstructions of aortic anatomy in a wide range of subjects

After validating the ability to assess aortic miscannulation rates in elderly cadavers using the electronic simulator, we proceeded to analyze a broader patient population. Following Institutional Review Board approval, a trauma database was searched to identify a total of *n*=89 thin-section, contrast-enhanced chest-abdomen-pelvis CTAs from 5-93-year-old subjects. Trauma patients were used because they covered a wide age range. Mean age was 47±24 years, and 10 subjects represented most decades (except first n=6 and 6th n=13). The majority of subjects were male (71%). Scans were obtained with a GE LightSpeed VCT XT 64-channel CTA scanner (GE Healthcare, Waukesha, USA) with a slice thickness of 1.25 mm and a resolution of 512 × 512 pixels. Patients were injected with 75 mL intravenous contrast delivered at 3 mL/s.

Similar to reconstructions in cadavers, 3D vascular anatomy (Figure 1) included the entire aorta and the common carotid, subclavian, axillary, celiac, superior mesenteric, renal, iliac, and common femoral arteries^7,8^. Interobserver variability was assessed using a training dataset of 5 randomly selected scans. Each reconstruction was overlaid onto the baseline image to quantify the differences at 10 random locations^7^. The training was considered satisfactory when the cumulative error was <5% in all locations.

**Figure 1.**
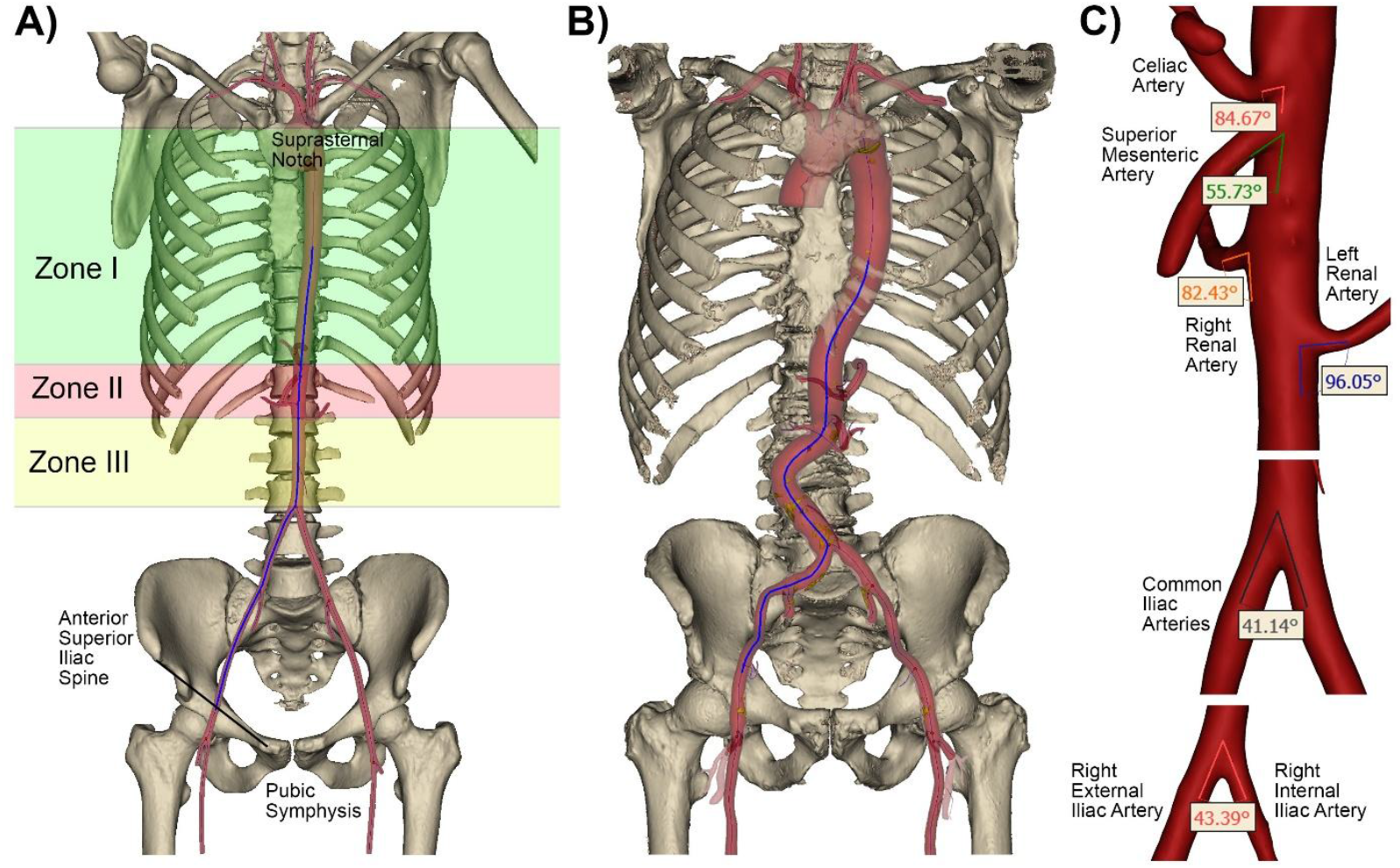
Three-dimensional reconstructions of vasculature and bones in representative young (A) and old (B) subjects. Aortic zones are marked with green, red, and yellow colors. The arterial centerline from the femoral access site to the center of Zone I^11^ is blue. C) Measurements of branch angles along the catheter’s path from the femoral access site to the center of aortic Zone I.

After completing three-dimensional reconstructions, centerlines were fit into the arterial volumes and used to measure tortuosity and distance from the femoral artery access site located between the pubic symphysis and the anterior superior iliac spine to the center of aortic Zone I^11^ located between the left subclavian and the celiac arteries (Figure 1A, B). Additionally, tortuosity was measured separately in two segments – from the femoral access site to the common iliac artery bifurcation and from the common iliac artery bifurcation to the center of aortic Zone I^11^. We defined tortuosity as a fractional increase in the length of the centerline of the tortuous vessel relative to a perfectly straight path, i.e. (distance along the centerline – straight distance)/distance along the centerline. Branching angles were measured in 3D between the external and the internal iliac arteries, common iliac arteries, renal arteries and the aorta, and superior mesenteric and celiac arteries and the aorta (Figure 1C). For consistency, angles between the renal, superior mesenteric, celiac arteries, and the aorta were measured using the distal aspect of the aorta as demonstrated in Figure 1, so angles >90° indicated that arteries pointed cephalad. Torso length was measured as the shortest distance between the pubic symphysis and the suprasternal notch.

### 2.5 Assessment of aortic miscannulation frequency using an electronic simulator

Patient-specific anatomies reconstructed in Mimics were exported in STL format and loaded into the VIST G5 simulator using the CASE-IT EVAR module, following the same procedure used for cadaveric vascular anatomies. Each anatomy was also mapped to the template reference points to ensure the proper location of arteries and other anatomic structures, and the same 0.035” 35° angle hydrophilic guidewire was selected for navigation. An electric automatic wire-feeding mechanism was again used to advance a physical guidewire into the simulator using 12 trials for each anatomy for a total of 2,136 guidewire passages. Final locations of the guidewire tip were recorded, and the percentage of unsuccessful attempts that resulted in the wire tip ending outside of the aorta (i.e., in a smaller artery) was calculated.

### 2.6 Statistical analysis

Correlations between variables were assessed using Pearson’s correlation *r*, and the hypothesis of no correlation was tested against the alternative that there is a nonzero correlation assuming statistical significance at p<0.05. When appropriate, multiple linear regression analysis was performed with SPSS v25 (IBM, Armonk, New York). Stepwise linear regression was used to determine statistically significant predictors. A variable was entered into the model when the significance level of its F value was less than 0.05, and the variable was removed from the model when the significance level was above 0.10. Model quality was assessed with adjusted R^2^.

## RESULTS

### 3.1 Aortic miscannulation rates in human cadavers compared with the electronic simulator

Aortic miscannulation in cadavers occurred in 7 out of 60 wire passes, i.e., at 11.7% frequency, and the most common final location for the miscannulated wire was either the contralateral or the ipsilateral iliac artery. Of these 7 miscannulations, 5 occurred in one anatomy, so that a total of 30% of subjects had at least one aortic miscannulation during multiple wire passage attempts. When using the same anatomies in Mentice VIST G5, the aortic miscannulation rate was exactly the same, i.e., 7 (11.7%) out of 60 wire passage attempts, but the miscannulations occurred in 4 instead of 3 subjects. Of these 4 subjects, 3 were the same cadaveric anatomies that had miscannulation in the cadaver lab, and the wire misplacements were also either into the contralateral or to the ipsilateral iliac artery. These results confirmed that the electronic simulator provided an accurate assessment of aortic miscannulation frequency when using the selected guidewire and could be used for the analysis of larger anatomic datasets.

### 3.2 Anatomic variation as a function of age

Measurements of the centerline distance from the femoral artery access site to the center of the aortic Zone I for all age groups are summarized in Figure 2A. Age and sex explained 37% of the variation in this distance, with age having a stronger effect than sex. The equation predicting distance from the femoral access site to the center of Zone I (mm) was 345+1.09·Age (years) + 24.78·Sex (1=Male/0=Female). Torso length and age explained 83% of the variation in the same centerline distance, with the equation 38+0.66·Torso length (mm) + 0.67·Age (years). Torso length had a significantly stronger effect than age (standardized β=0.75 versus β=0.34) and appeared to absorb the statistically significant effect of sex.

**Figure 2.**
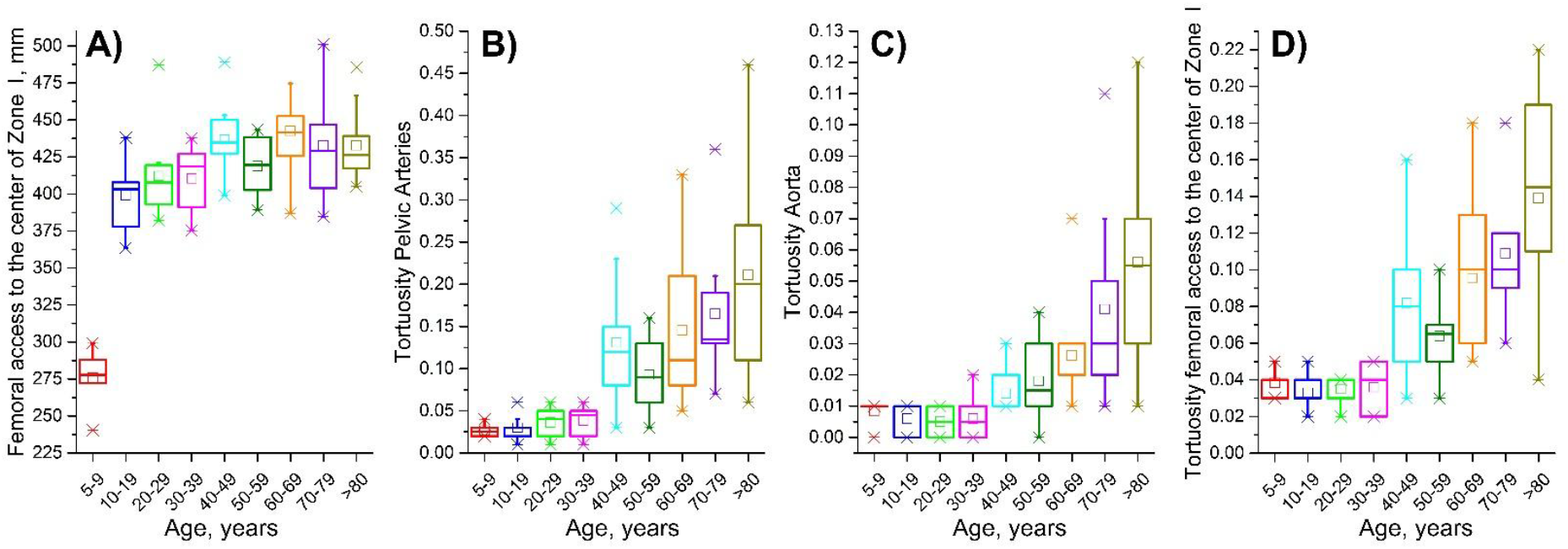
A) distance from the femoral access site to the center of aortic Zone I measured in mm along the arterial centerline, and tortuosity of B) pelvic arteries (i.e., from the femoral artery access site to the common iliac artery bifurcation), C) the aorta (i.e., from the common iliac artery bifurcation to the center of aortic Zone I), and D) total tortuosity measured from the femoral artery access site to the center of aortic Zone I. Box plots represent different age groups and boxes bound 25th and 75th percentiles, whiskers extend to 5th and 95th percentiles, 99th percentile is marked with a cross (x), and maximum values are represented by a minus sign (-). Additionally, mean values are marked with a hollow square, and a horizontal line within each box represents the median.

Age had a significant positive effect on tortuosity, demonstrating more tortuous arteries in older subjects (Figure 2 B, C, D). Pearson correlations between age and tortuosity of the pelvic arteries from the femoral artery access site to the common iliac artery bifurcation were *r*=0.70 (p<0.01), and for the aortic segment from the common iliac artery to the center of Zone I *r*=0.66 (p<0.01). The correlation between age and the overall tortuosity from the femoral artery access site to the center of aortic Zone I was *r*=0.73 (p<0.01).

The aortoiliac bifurcation angle widened with age (*r=*0.22, p=0.04), and the angle between the aorta and the celiac artery decreased with age (*r*=-0.22, p=0.04). Angles between the aorta and the internal iliac artery (p=0.65), renal arteries (p=0.18 and p=0.15), and the superior mesenteric artery (p=0.22) did not change significantly with age. Average angles for all measured arteries are summarized in Table 1, along with their standard deviations and the 5^th^ and 95^th^ percentile ranges. The angle between the left renal artery and the aorta was larger than that between the right renal artery and the aorta (p<0.01) by an average of 10°. A total of 11% of right renal arteries, 27% of left renal arteries, and 18% of superior mesenteric and 19% of celiac arteries pointed cephalad, i.e., the angle between the artery and the distal part of the aorta was >90°.

**Table 1.**
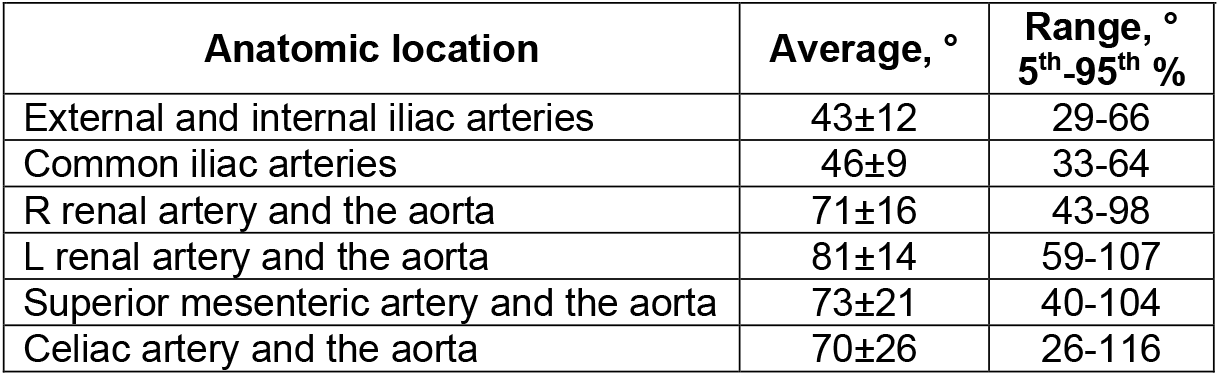
Average arterial angles (°), their standard deviations, and range between 5^th^ and 95^th^ percentiles. Note that angles >90° for the renal, superior mesenteric, and celiac arteries indicate that the artery is pointed cephalad.

| Anatomic location | Average, $^{\circ}$ | Range, $^{\circ}$<br>5<sup>th</sup>-95<sup>th</sup> % |
| --- | --- | --- |
| External and internal iliac arteries | 43±12 | 29-66 |
| Common iliac arteries | 46±9 | 33-64 |
| R renal artery and the aorta | 71±16 | 43-98 |
| L renal artery and the aorta | 81±14 | 59-107 |
| Superior mesenteric artery and the aorta | 73±21 | 40-104 |
| Celiac artery and the aorta | 70±26 | 26-116 |

### 3.3 Aortic miscannulation rates in a wide range of anatomies assessed using the electronic simulator

Aortic miscannulation frequency (%) plotted as a function of age is presented in Figure 3 and Figure 4. Figure 3 demonstrates the frequency of at least one aortic miscannulation in 12 trials for each anatomy, and Figure 4 presents miscannulation frequency as a function of all attempts. Overall, wire advancement was successful in 64% of anatomies in all 12 attempts, and miscannulation occurring at least once was seen in up to 60% of subjects in a given age group.

**Figure 3.**
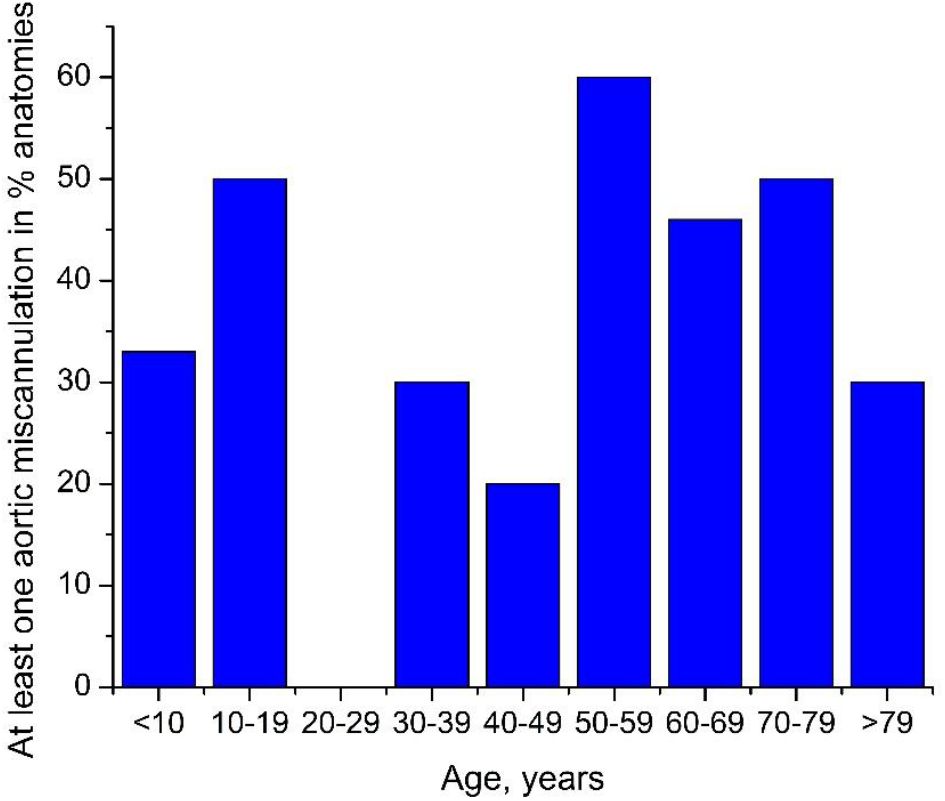
Frequency of at least one aortic miscannulation in anatomies of different ages.

**Figure 4.**
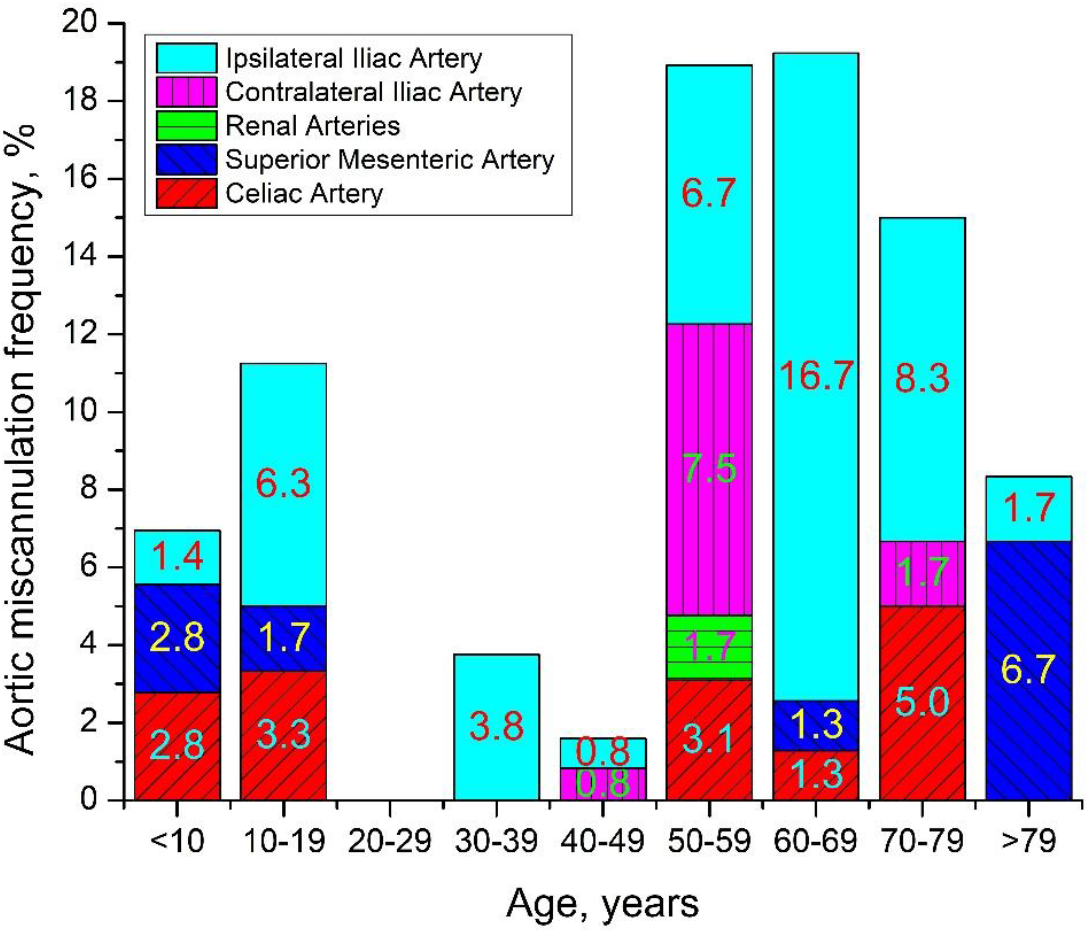
Aortic miscannulation frequency in different age groups.

The average frequency of aortic miscannulation was 9.9±17.7% when counting all wire passage attempts. In subjects younger than 50 years, aortic miscannulation occurred in 4.5±10.6%, while in subjects older than 50 years, miscannulations were more common (p<0.01) and occurred in 15.6±21.8% of wire passages. This is comparable to the 11.7% miscannulation rate observed in older cadavers. Figure 4 demonstrates that the most frequent aortic miscannulations occurred in the ipsilateral internal iliac artery that, on average, was observed in 5.1% of all wire passages. The second most common miscannulation location was the celiac (1.7%) followed by the superior mesenteric arteries (1.4%).

## DISCUSSION

Malposition of REBOA balloons into the iliac arteries^15^ and higher complication rates of ECMO miscannulation without the assistance of fluoroscopy^6^ point to the importance of characterizing miscannulation rates during fluoroscopy-free endovascular navigation. Our study reported on the frequency of aortic miscannulation, which can inform the risk of performing fluoroscopy-free catheterization, help avoid associated clinical complications, inform patient selection, and guide the development of smarter devices for endovascular surgery. To quantify the miscannulation frequency in a wide range of anatomies, we have used an electronic simulator validated with human cadaver experiments. The main advantage of this approach is that it allows a cost-effective and rigorously controlled way of performing vascular navigation in a large number of complex vascular anatomies. Our data demonstrate that fluoroscopy-free aortic navigation may be associated with 5-19% rate of aortic miscannulation. The majority of miscannulations were into the iliac, celiac, and superior mesenteric arteries, and the significant size differences between these vessels and the aorta suggest that deployment of aortic-size devices, such as a stent-grafts or balloons, should be performed only after verifying their position to avoid complications such as vessel rupture that would be difficult to control^16^.

Aortic miscannulations were observed in all age groups but were particularly prevalent in subjects older than 50 years, with an overall miscannulation rate reaching 19%. Higher miscannulation rates in older anatomies are likely associated with increased vessel tortuosity^8,17–19^, larger aortic diameters^7^, and wider aortoiliac bifurcation angles^8^ that can produce dangerous scenarios for fluoroscopy-free navigation. Nevertheless, aortic miscannulations also occurred in younger subjects, likely because most vessel angulations did not appear to depend on age, and 10-30% of all renal, superior mesenteric, and celiac arteries were pointed cephalad. This suggests that the risk of miscannulation into a small vessel is sizable in subjects of any age, which points to the need for developing smarter devices or other adjunctive technologies to detect incorrect placement and prevent vessel injury, particularly in settings with limited imaging capabilities.

Centerline distances from the femoral access site to the center of aortic Zone I used in our study directly informed the inserted catheter lengths, but these distances are not easily obtained without imaging and are usually roughly estimated by torso length^20^. Morphometric maps^8^ allow for better length estimation in older subjects as uncorrected lengths can increase the likelihood of wire misplacement in the aortic Zone II when aiming for Zone I, or in a smaller artery (such as the common iliac artery) when aiming for the aortic Zone III^11^. These results highlight the importance of proper patient selection when evaluating the potential risks and benefits of fluoroscopy-free endovascular navigation and suggest that extra care should be taken when calculating insertion lengths in older subjects.

While our data provide an assessment of aortic miscannulation frequency in anatomies of different ages, it is important to be mindful of study limitations. First, most electronic simulators were designed for educational purposes^21–26^ and therefore may not simulate the entire complexity of human vascular biomechanics and artery-device interaction. While we have validated our approach using human cadavers, these also have their limitations related to older age and stiffer vessels compared to younger patients. Second, in our assessments, we have used a regular stiffness regular angled guidewire, which is only one of the many available wires. Other devices may have different characteristics and may result in different miscannulation rates, which warrants further studies. Lastly, while the analyzed sample size (n=89) was significant, a greater number of vascular anatomies may provide additional refinement in the observed miscannulation rates as it would allow encountering more anatomical variations. Despite these limitations, the current study represents an initial and essential first step towards understanding how to mitigate aortic miscannulation during fluoroscopy-free endovascular navigation in patient populations of varying ages while highlighting the importance of the continued advancement of endovascular device technology.

## Data Availability

All data produced in the present study are available upon reasonable request to the authors

## COI/DISCLOSURES

The authors declare no relevant conflict of interest.

## FUNDING/FINANCIAL SUPPORT

The research reported in this publication was supported in part by the U.S. Army Medical Research and Materiel Command under award number W81XWH-16-2-0034 (Log 14361001) and by the National Heart, Lung, And Blood Institute of the National Institutes of Health under award numbers HL125736, HL147128, and P20GM152301.

## DATA STATEMENT

Data can be available from the corresponding author upon a reasonable request.

## APPENDIX

### Phantom anatomies to validate the Mentice VIST G5 simulator

Phantom geometries (Figure A1) contained straight (Figure A1 A, B) and curved bifurcating tubes (Figure A1 C, D) of different diameters to test the simulator’s ability to register inserted wire lengths and evaluate how the wire traversed straight and curved anatomies.

**Figure A1.**
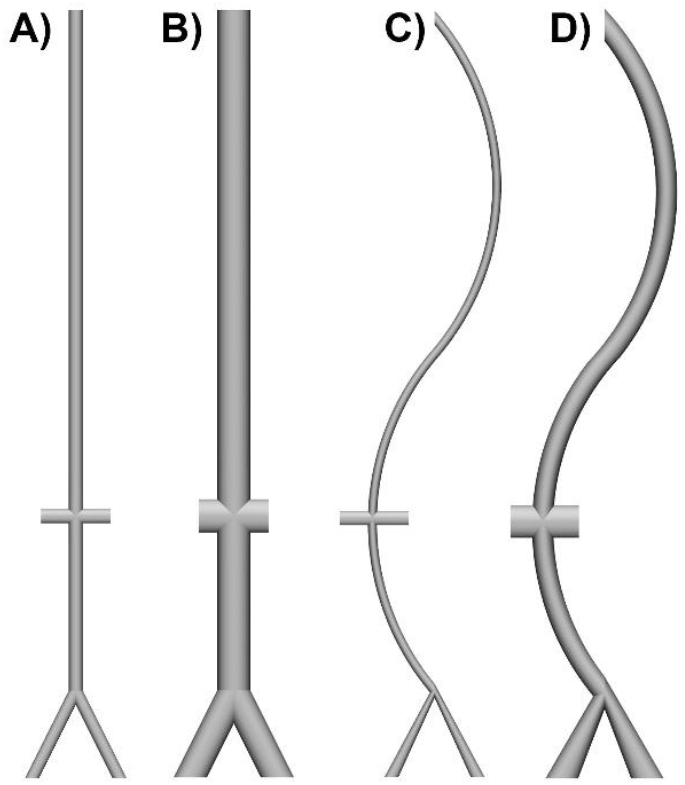
Phantom geometries used in the VIST G5 simulator to validate insertion lengths, accuracy, and repeatability of measurements.

### The automated wire feeding system

To ensure maximal accuracy and consistency between wire advancements, a wire feeding system was developed that contained a 24V Wire Feed Assembly Motor and a DC power source. Voltage varied from 2 to 8V, which produced different wire feeding rates. The prescribed length of wire, consistent with the length of the phantom anatomy, was then advanced using the automatic wire-feeder, and the wire travel distance was measured in the simulator and compared with the actual advancement distance. Testing at each voltage level was repeated several times to assess consistency.

Results demonstrated that the optical wire tracking system of the VIST G5 was sensitive to the wire advancement rate. Figure A2 illustrates different wire lengths required to cover 610mm distance in the electronic simulator plotted as a function of DC motor supply, i.e., the rate of wire advancement. Measurements were done in a thick straight phantom anatomy B plotted in Figure A1. Results demonstrate that voltage >4V resulted in poor wire tracking and a large discrepancy in measurements. Therefore all measurements in patient-specific anatomies were performed at 2V DC power (corresponded to 18.4 mm/s linear wire advancement rate), which ensured maximum accuracy and consistency of measurements.

**Figure A2.**
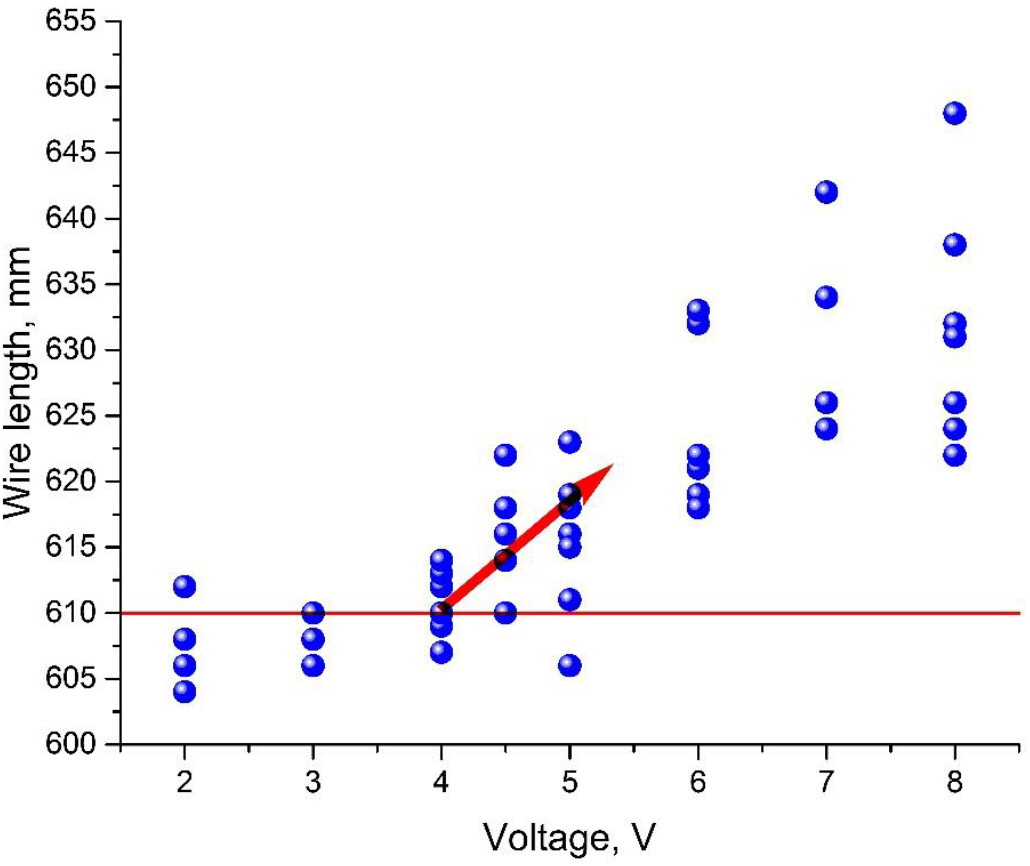
Wire length required to reach 610mm mark in the electronic simulator as a function of DC power supply (V) to the automatic wire-feeder. Measurements were performed in a thick straight phantom anatomy B plotted in Figure A1. Note higher variability at a supplied voltage over 4V, likely due to inconsistent optical tracking of the wire at higher advancement speeds.

## Notes

### Competing Interest Statement

The authors have declared no competing interest.

### Author Declarations

The Institutional Review Board of the University of Nebraska Medical Center gave ethical approval for this work involving analysis of human CT angiography data (IRB #0211-13-EX). Research involving human cadavers was conducted under institutional policies governing the use of anatomical materials; because cadavers are not considered human subjects, the Institutional Review Board of the University of Nebraska Medical Center waived ethical approval for this work.

## REFERENCES

1. Writing Committee. Editor’s Choice – Management of Descending Thoracic Aorta Diseases: Clinical Practice Guidelines of the European Society for Vascular Surgery (ESVS). Eur J Vasc Endovasc Surg. 2017;53(1):4–52. doi:10.1016/j.ejvs.2016.06.005

2. Nienaber CA, Fattori R, Lund G, et al. Nonsurgical Reconstruction of Thoracic Aortic Dissection by Stent–Graft Placement. N Engl J Med. 1999;340(20):1539–1545. doi:10.1056/NEJM199905203402003

3. Xenos ES, Abedi NN, Davenport DL, et al. Meta-analysis of endovascular vs open repair for traumatic descending thoracic aortic rupture. J Vasc Surg. 2008;48(5):1343–1351. doi:10.1016/j.jvs.2008.04.060

4. Reuben BC, Whitten MG, Sarfati M, Kraiss LW. Increasing use of endovascular therapy in acute arterial injuries: analysis of the National Trauma Data Bank. J Vasc Surg. 2007;46(6):1222–1226. doi:10.1016/j.jvs.2007.08.023

5. Kato N, Dake MD, Miller DC, et al. Traumatic thoracic aortic aneurysm: treatment with endovascular stent-grafts. Radiology. 1997;205(3):657–662. doi:10.1148/radiology.205.3.9393517

6. Kashiura M, Sugiyama K, Tanabe T, Akashi A, Hamabe Y. Effect of ultrasonography and fluoroscopic guidance on the incidence of complications of cannulation in extracorporeal cardiopulmonary resuscitation in out-of-hospital cardiac arrest: A retrospective observational study. BMC Anesthesiol. 2017;17(1):1–7. doi:10.1186/s12871-016-0293-z

7. Kamenskiy A, Miserlis D, Adamson P, et al. Patient demographics and cardiovascular risk factors differentially influence geometric remodeling of the aorta compared with the peripheral arteries. Surgery. 2015;158(6):1617–1627. doi:10.1016/j.surg.2015.05.013

8. MacTaggart JN, Poulson WE, Akhter M, et al. Morphometric roadmaps to improve accurate device delivery for fluoroscopy-free resuscitative endovascular balloon occlusion of the aorta. J Trauma Acute Care Surg. 2016;80(6):941–946. doi:10.1097/TA.0000000000001043

9. Evans C, Schlitzkus L, Schiller A, Kamenskiy A, MacTaggart J. Comparison of Simulation Models for Training a Diverse Audience to Perform Resuscitative Endovascular Balloon Occlusion of the Aorta. J Endovasc Resusc Trauma Manag. 2019;3(3). doi:10.26676/jevtm.v3i3.86

10. Wadman MC, Lomneth CS, Hoffman LH, Zeger WG, Lander L, Walker R a. Assessment of a new model for femoral ultrasound-guided central venous access procedural training: a pilot study. Acad Emerg Med. 2010;17(1):88–92. doi:10.1111/j.1553-2712.2009.00626.x

11. Stannard A, Eliason JL, Rasmussen TE. Resuscitative endovascular balloon occlusion of the aorta (REBOA) as an adjunct for hemorrhagic shock. J Trauma. 2011;71(6):1869–1872. doi:10.1097/TA.0b013e31823fe90c

12. Lombardi J V., Hughes GC, Appoo JJ, et al. Society for Vascular Surgery (SVS) and Society of Thoracic Surgeons (STS) reporting standards for type B aortic dissections. J Vasc Surg. 2020;71(3):723–747. doi:10.1016/j.jvs.2019.11.013

13. Kamenskiy A, Aylward P, Desyatova A, DeVries M, Wichman C, MacTaggart J. Endovascular Repair of Blunt Thoracic Aortic Trauma is Associated With Increased Left Ventricular Mass, Hypertension, and Off-target Aortic Remodeling. Ann Surg. 2020;XX(Xx):1. doi:10.1097/SLA.0000000000003768

14. Jadidi M, Poulson W, Aylward P, et al. Calcification prevalence in different vascular zones and its association with demographics, risk factors, and morphometry. Am J Physiol Heart Circ Physiol. 2021;320(6):H2313–H2323. doi:10.1152/ajpheart.00040.2021

15. Davidson AJ, Russo RM, Reva VA, et al. The pitfalls of resuscitative endovascular balloon occlusion of the aorta: Risk factors and mitigation strategies. J Trauma Acute Care Surg. 2018;84(1):192–202. doi:10.1097/TA.0000000000001711

16. Skitch S, Engels PT. Acute Management of the Traumatically Injured Pelvis. Emerg Med Clin North Am. 2018;36(1):161–179. doi:10.1016/j.emc.2017.08.011

17. Tawfik AM, Sobh DM, Gadelhak B, Sobh HM, Batouty NM. The effect of age and gender on tortuosity of the descending thoracic Aorta. Eur J Radiol. 2019;110(November 2018):54–59. doi:10.1016/j.ejrad.2018.11.017

18. Sugawara J, Hayashi K, Yokoi T, Tanaka H. Age-associated elongation of the ascending aorta in adults. JACC Cardiovasc Imaging. 2008;1(6):739–748. doi:10.1016/j.jcmg.2008.06.010

19. Wolf YG, Tillich M, Lee W a, Rubin GD, Fogarty TJ, Zarins CK. Impact of aortoiliac tortuosity on endovascular repair of abdominal aortic aneurysms: evaluation of 3D computer-based assessment. J Vasc Surg. 2001;34(4):594–599. doi:10.1067/mva.2001.118586

20. Stannard A, Morrison JJ, Sharon DJ, Eliason JL, Rasmussen TE. Morphometric analysis of torso arterial anatomy with implications for resuscitative aortic occlusion. J Trauma Acute Care Surg. 2013;75(2):S169–S172. doi:10.1097/TA.0b013e31829a098d

21. Eslahpazir BA, Goldstone J, Allemang MT, Wang JC, Kashyap VS. Principal considerations for the contemporary high-fidelity endovascular simulator design used in training and evaluation. J Vasc Surg. 2014;59(4):1154–1162. doi:10.1016/j.jvs.2013.11.074

22. Willaert WIM, Aggarwal R, Herzeele I Van, Cheshire NJ, Vermassen FE. Recent advancements in medical simulation: Patient-specific virtual reality simulation. World J Surg. 2012;36:1703–1712. doi:10.1007/s00268-012-1489-0

23. Cheng I, Shen R, Moreau R, Brizzi V, Rossol N, Basu A. An augmented reality framework for optimization of computer assisted navigation in endovascular surgery. 2014 36th Annu Int Conf IEEE Eng Med Biol Soc EMBC 2014. 2014;1:5647–5650. doi:10.1109/EMBC.2014.6944908

24. Chaer RA, DeRubertis BG, Lin SC, et al. Simulation improves resident performance in catheter-based intervention: Results of a randomized, controlled study. Ann Surg. 2006;244(3):343–349. doi:10.1097/01.sla.0000234932.88487.75

25. Dawson DL, Lee ES, Hedayati N, Pevec WC. Four-Year Experience with a Regional Program Providing Simulation-Based Endovascular Training for Vascular Surgery Fellows. J Surg Educ. 2009;66(6):330–335. doi:10.1016/j.jsurg.2009.07.004

26. Dawson DL, Meyer J, Lee ES, Pevec WC. Training with simulation improves residents’ endovascular procedure skills. J Vasc Surg. 2007;45(1):149–154. doi:10.1016/j.jvs.2006.09.003

